# Molecular Detection of Human Metapneumovirus in Lagos, Nigeria

**DOI:** 10.1101/2025.02.21.25322649

**Authors:** Abdul-Azeez Adeyemi Anjorin, Haminat Oluwatoyosi Olaleye, Saburi Adekilekun Sayid, Tomisin Deborah Ajose, Grace Treasure Daniel, Kabir Olusegun Akinyemi

## Abstract

**Background:** Human metapneumovirus (HMPV) is an important respiratory pathogen worldwide. It is a leading cause of lower respiratory infection in infants, children, elderly and immunocompromised individuals.

**Methods:** This cross-sectional molecular epidemiological study examined HMPV among people with underlying conditions in different clinics in Lagos. Nasopharyngeal samples collected from 150 patients aged 1-88 years between May and June, 2022 were tested by Real-Time Polymerase Chain Reaction. Samples were extracted using Addbio extraction kit (Germany) while master-mix was prepared with primer designed targeting the F protein gene.

**Results:** A total of 15.3% (23/150) was positive for HMPV. The highest HMPV molecular prevalence of 34.8% (8/23) was recorded among the age group (>55 years) while the lowest prevalence of 4.3% (1/23) was recorded in the age group (12-22 years). Male predominance of 56.6% (13/23) was chronicled. Interestingly, HMPV molecular prevalence of 43.3% (10/23), 26.1% (6/23), 21.7% (5/23), 17.4%, (4/23), 13.3% (3/23) were found with co-infection/co-morbidity in malaria, high blood pressure, diabetes, tuberculosis, and pneumonia respectively. Based on location, the highest molecular prevalence was recorded in Badagry 39.1% (9/23), followed by Ojo 34.8% (8/23), while Alimosho recorded the lowest prevalence of 26.1% (6/23). Traders had the highest 39.1% (9/23) molecular prevalence while the lowest 4.3% (1/23) was recorded among patients that are civil servants, musicians, teachers and the unemployed.

**Conclusion:** The interaction between HMPV and other pathogens, such as malaria and respiratory viruses suggests that HMPV may exacerbate existing health conditions or that individuals with these conditions are more prone to HMPV infection. This highlights the need for comprehensive diagnostic approaches in clinical settings to identify and manage multiple infections effectively. The findings accentuates the importance of epidemiological surveillance especially among individuals having underlying diseases and protracted illnesses.

## 1. INTRODUCTION

The most common disease in the world are the respiratory infections (Monto *et al*., 2002) and they are responsible for considerable morbidity and mortality especially in infants and elderly (Nicholas *et al*., 2020). Human metapneumovirus (HMPV) is a major worldwide respiratory pathogen which can both be symptomatic and asymptomatic in infected host (Bruno *et al*., 2009). It was first isolated in Netherlands from children with respiratory syncytial virus-like symptoms in previously virus-negative nasopharyngeal aspirates in the year 2001 (Hall, 2001; Van den Hoogen *et al*., 2001; Hoogen 2002). Although, Owor *et al*. (2016) documented its antibodies in patient serum dating back to the 1950s, indicating that the virus has been circulating undetected in the human population for decades, HMPV has also been detected in specimens from adults, elderly and immunocompromised patients suffering from acute respiratory tract infections (Boivin and Pelletier, 2002; Owor *et al*., 2016).

The transmission of HMPV is likely to occur by direct or indirect contact. It can be spread from an infected person to others through secretions from coughing and sneezing, close personal contacts, and touching the mouth, nose or eyes after touching objects or surfaces that have been contaminated with the virus. The incubation period is 3-6 days with a median duration of the illness that can vary depending upon severity (CDC, 2016). The clinical presentation usually erupts from common upper respiratory tract symptoms to severe pneumonia leading to diverse pathologies of the upper or lower respiratory tracts including laryngitis, rhinopharyngitis, pneumonia, croup, bronchiolitis or asthma (Biovin *et al*., 2003; Boivin *et al*., 2010). Although, the clinical course usually commences with an asymptomatic state that can last about seven (7) days post-exposure, trailed by a week symptoms of upper respiratory tract infection before the disease resolution. However, the clinical course can spread towards the lower respiratory tract resulting in the involvement of the lungs parenchymal and further complications (Boivin *et al*., 2010).

The ICTV 2021 classified HMPV under the realm Riboviria, kingdom Orthornavirae, and phylum known as Negarnaviricota. It belongs to the family Paramyxoviridae, sub family Pneumovirine, class Monjiviricetes, with the order Mononegavirales, and categorized as the first human member of the metapneumovirus genus. This family also contain measles, mumps, human parainfluenza viruses, and respiratory syncytial virus (RSV) (Haas *et al*., 2013).

In Africa, Human metapneumovirus associated acute respiratory tract infection (ARTI) has been reported in children (Divarathna *et al*., 2019). Oketch *et al*. (2021) investigation in children within the age range of 5 years documented 39 incidences in Mali and Zambia. Another 22.2% incidences were reported from Algeria by Derrar *et al.,* 2019 among 117 children by RT-PCR. A total of 13.8% incidence was reported among 295 children between 2-59 months after a multiplex RT-PCR in Madagascar (Hoffmann *et al.,* 2012). Jroundi *et al*. (2016) documented 8.9% incidence among 683 children > 5 years in Morocco. In Nigeria, Akinloye (2011) documented 3.7% prevalence of HMPV among children in Oyo State.

In St. Louis, Missouri, the predominant genotype of HMPV switched in consecutive years from genotype A to genotype B. Wang *et al* (2018) reported that HMPV was detected in 14.2 million ALRI cases, 643,000 hospital admissions, 7,700 in–hospital deaths, and 16,100 overall ALRI deaths among children under five years globally. Although, there have been many advances made over the years since its discovery, however there are supportive regiments that can be used.

Yet there are still no approved antivirals or vaccines available to treat with the dangerous sporadic resultant outbreaks including previous and current spread of HMPV in Asia which if not properly manage may spread to other parts of the world, hence we therefore investigated and herein report our findings on the presence of Human metapneumovirus in the general population in different communities in Lagos, Nigeria.

## 2. MATERIALS AND METHODS

### STUDY DESIGN AND LOCATION

This study is a descriptive, cross sectional, hospital-based molecular epidemiological surveillance in three (3) communities of Alimosho, Badagry and Ojo in Lagos, Nigeria. Lagos State with a population of over 21 million (21,000,000) and a total area of 3,577 square meters is located in the Southwestern part of Nigeria along the Atlantic Ocean with an average temperature of 26.7°C and annual rainfall of 178.3mm/70.2 inch. Lagos State is bounded by the Ogun State to the north and east, and by the inlet of Benin to the south, and to the West by the Republic of Benin. Specifically, samples were collected from Alimosho general hospital located in Igando, Bola Ahmed Tinubu health center located in Egbeda and Ipinilere primary health center located in Akowonjo. The hospital and centers provide 24 hours in-patients and out-patients supports and care for an average of 30,000 patients monthly.

Nasopharyngeal samples were collected from patients from the different clinics and hospitals by purposive sampling during the peak rainy period from June to July, 2022. Consent forms for the participants along with a pretested paper-interviewer questionnaires were appropriately administered by the research team members.

### INCLUSION AND EXCLUSION CRITERIA

The inclusion criteria were patients with symptoms similar to HMPV including fever, cough, dyspnoea, sputum production, and wheezing especially in young children and aged adults. The exclusion criteria were patients who have had COVID and those who didn’t give their consents.

### ETHICAL APPROVAL AND PERMISSION

Permission was sought after from the Heads of Health Centers where the samples were obtained while the ethical approval was obtained from LASUTH (Lagos State University Teaching Hospital) with the registration number NHREC04/04/2008 and reference number LREC/06/10/1272, respectively.

### DATA COLLECTION AND STATISTICAL ANALYSIS

The data were collected using a standard pretested questionnaire in which biodata (age, gender, occupation, educational level, marital status, place of origin) and clinical data (signs and symptoms) pathognomonic of HMPV such as fever, sore throat, cough, nausea, shortness of breath, wheezing, stuffed nose, tightness of chest were appropriately captured and documented (Fig. 1). Data obtained were entered into a spreadsheet database prepared with Microsoft excel. They were compared and changed for wrongful entries. The data were then sorted, averaged and organized into table using descriptive statistical tools.

**Fig 1.**
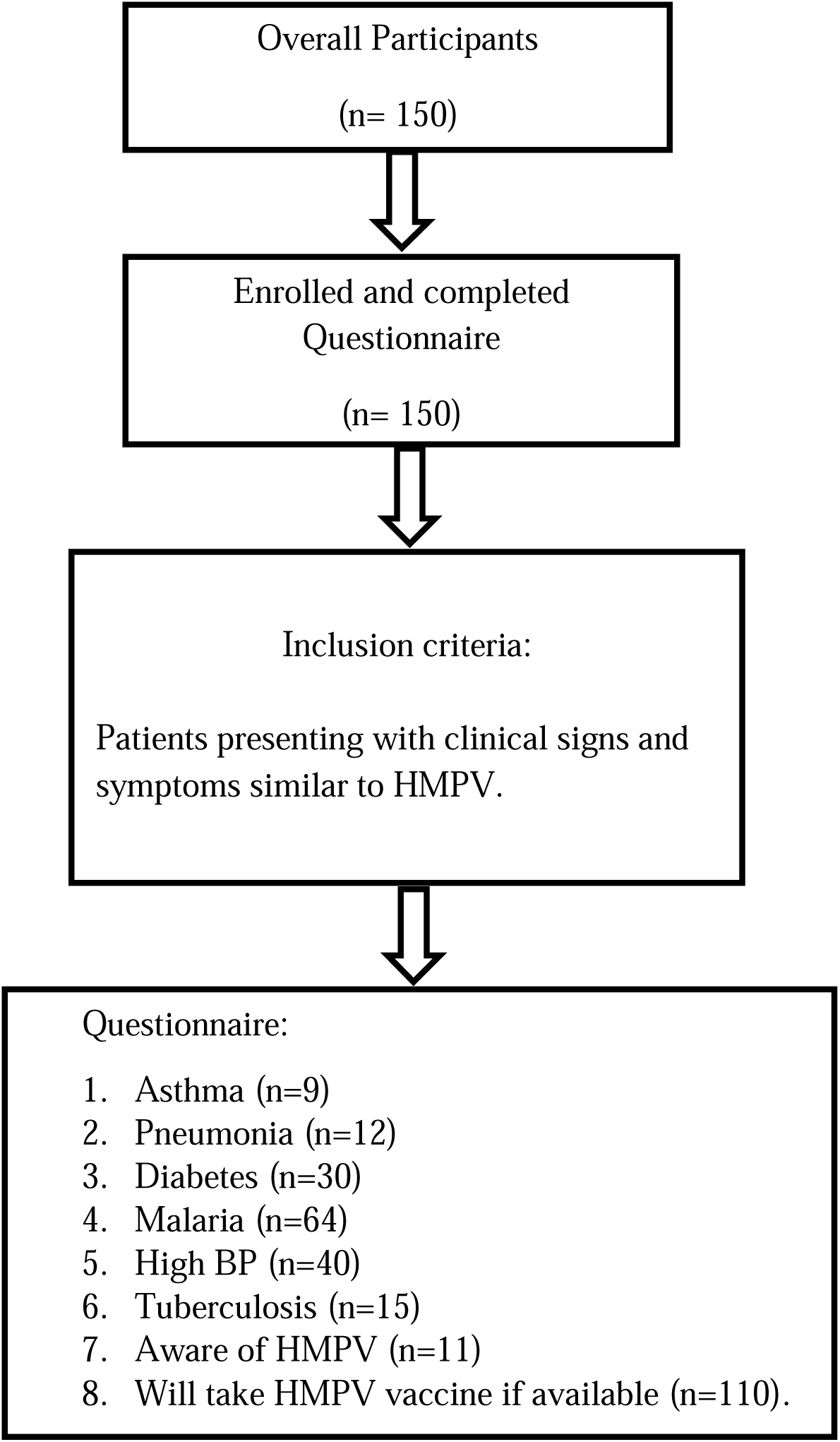
Flow chart of inclusion criteria and distribution of clinical signs and symptoms.

### LABORATORY ANALYSIS

The study laboratory analyses were carried out at the Virology Laboratory, Department of Microbiology Molecular Research Lab, Faculty of Science, Lagos State University, Ojo.

### SAMPLE COLLECTION AND TESTING

A total of 150 nasopharyngeal samples were collected using Dracon swabs with flexible plastic shafts. The swab was inserted gently and slowly through the nostril parallel to the palate, rubbed and rolled to absorb secretions collected from both nostrils using the same swab. This was then transferred into a cryovial containing viral transport medium (VTM). Each cryovial was labelled with unique laboratory code number and were appropriately transported in cold chain to the laboratory for freezing at -30 until analysis.

#### Extraction process

Equal aliquots of three (3) human nasal samples were pooled into one cryovial tube to achieve the given 200µl needed for extraction. A total of 150 samples were pooled into 50 cyrovial tubes. Nucleic acid was extracted with AddPrep Total RNA extraction kit (Addbio, Sweden) following the manufacturer’s instructions. Accurately measured 200µl each of the sample aliquots was dispensed in a microcentrifuge tube followed by the addition of 350µl lysis buffer with pre-added beta-mercaptoethanol in order to neutralize ribonucleases released from the viral entity. This was followed by vortexing and incubation at room temperature for 10 minutes. A 150µl aliquot of isopropanol was added to obtain each sample lysate. Each lysate was vortexed and transferred into a spin column. The mixture was centrifuged at 13,000 rpm for 1 minute while the flow-through was discarded and collection tube was blotted on sterile tissue paper. A 500µl of wash-1 buffer was then added to the spin column, centrifuged at 13,000 rpm for 1 minute, then flow-through was also discarded and collection tube was blotted on sterile tissue paper. The same procedure was taken for wash-2 buffer. The spin column was centrifuged again at 13,000 rpm to remove the residual ethanol. The spin column was then placed into a new nuclease free microcentrifuge tube. Then 100µl elution buffer was added to the center of the column, it was incubated at room temperature for two minutes. Final centrifugation was carried out at 13,000 rpm for 1 minute to elute the RNA. The supernatant was separated and appropriately stored in Eppendorf tube before amplification the next day.

#### Master-mixture and RT-qPCR Amplification

Real-time PCR master mixture was performed using Luna Universal One-Step RT-qPCR kits (BioLabs, UK) following manufacturer’s instructions. A total of 0.8 µl forward and reverse primer (F-HMPV 5-GCC GTT AGC TTC AGT CAA TTC AA-3; and reverse-HMPV 5-TCC AGC ATT GTC TCT GAA AAT TGC-3) targeting the F protein gene was added to the aliquot, along with 5µl of nuclease-free water. PCR amplification was carried out in a Rotor gene thermocycler (Qiagen, Germany) following thermocycling conditions of reverse transcription at 55°C for 10 minutes (1 cycle), initial denaturation at 94°C for 10 minutes (1 cycle), denaturation at 94°C for 10 seconds (45 cycles), and final extension at 60°C for 30 seconds (45 cycles).

## 3. RESULTS

A total of 15.3% (23/150) were positive for HMPV by RT-qPCR. The findings showed data with different epidemiological parameters including age, gender, location, occupation and clinical presentations (Table 1). The highest HMPV prevalence of 34.8% (8/23) was recorded among the age group (>55 years) while the lowest prevalence 4.3% (1/23) was recorded in the age group (12-22 years). Based on gender distribution, males were examined to be more positive with a predominance of 56.6% (13/23). Interestingly, HMPV molecular prevalence of 43.3% (10/23), 26.1% (6/23), 21.7% (5/23), 17.4%, (4/23), and 13.3% (3/23) were found with co-infection/co-morbidity in malaria, high blood pressure, diabetes, tuberculosis, and pneumonia respectively. Based on location, the highest molecular prevalence was recorded in Badagry 39.1% (9/23), followed by Ojo 34.8% (8/23), while Alimosho recorded the lowest prevalence of 26.1% (6/23). Traders had the highest 39.1% (9/23) molecular prevalence while the lowest 4.3% (1/23) was recorded among patients that are civil servants, musicians, teachers and the unemployed.

**Table 1:** Epidemiological data of the general population with HMPV in Lagos, Nigeria.

| **Epidemiological data** | **Total (%)**<br>**(n=150)** | **HMPV Positive (%)**<br>**(n=23)** |
| --- | --- | --- |
| **Age Group** |  |  |
| *1-11* | 2 (1.3) | 0 (0) |
| *12-22* | 18 (12.0) | 1 (4.3) |
| *23-33* | 48 (32.0) | 5 (21.7) |
| *34-44* | 28 (18.7) | 3 (13.0) |
| *45-55* | 18 (12.0) | 6 (26.1) |
| *>55* | 35 (23.3) | 8 (34.8) |
| **Gender** |  |  |
| *Male* | 73 (48.7) | 13 (46.4) |
| *Female* | 77 (51.3) | 10 (35.7) |
| **Location** |  |  |
| *Alimosho* | 50 (33.3) | 6 (26.1) |
| *Badagry* | 50 (33.3) | 9 (39.1) |
| *Ojo* | 50 (33.3) | 8 (34.8) |
| **Clinical presentation** |  |  |
| *Asthma* | 10 (6.7) | 0(0) |
| *Diabetes* | 31 (20.5) | 5 (21.7) |
| *High BP* | 40 (26.7) | 6 (26.1) |
| *Malaria* | 66 (44.0) | 10 (43.5) |
| *Tuberculosis* | 16 (10.7) | 4 (17.4) |
| *Pneumonia* | 16 (10.7) | 3 (13.0) |

**Fig. 2:**
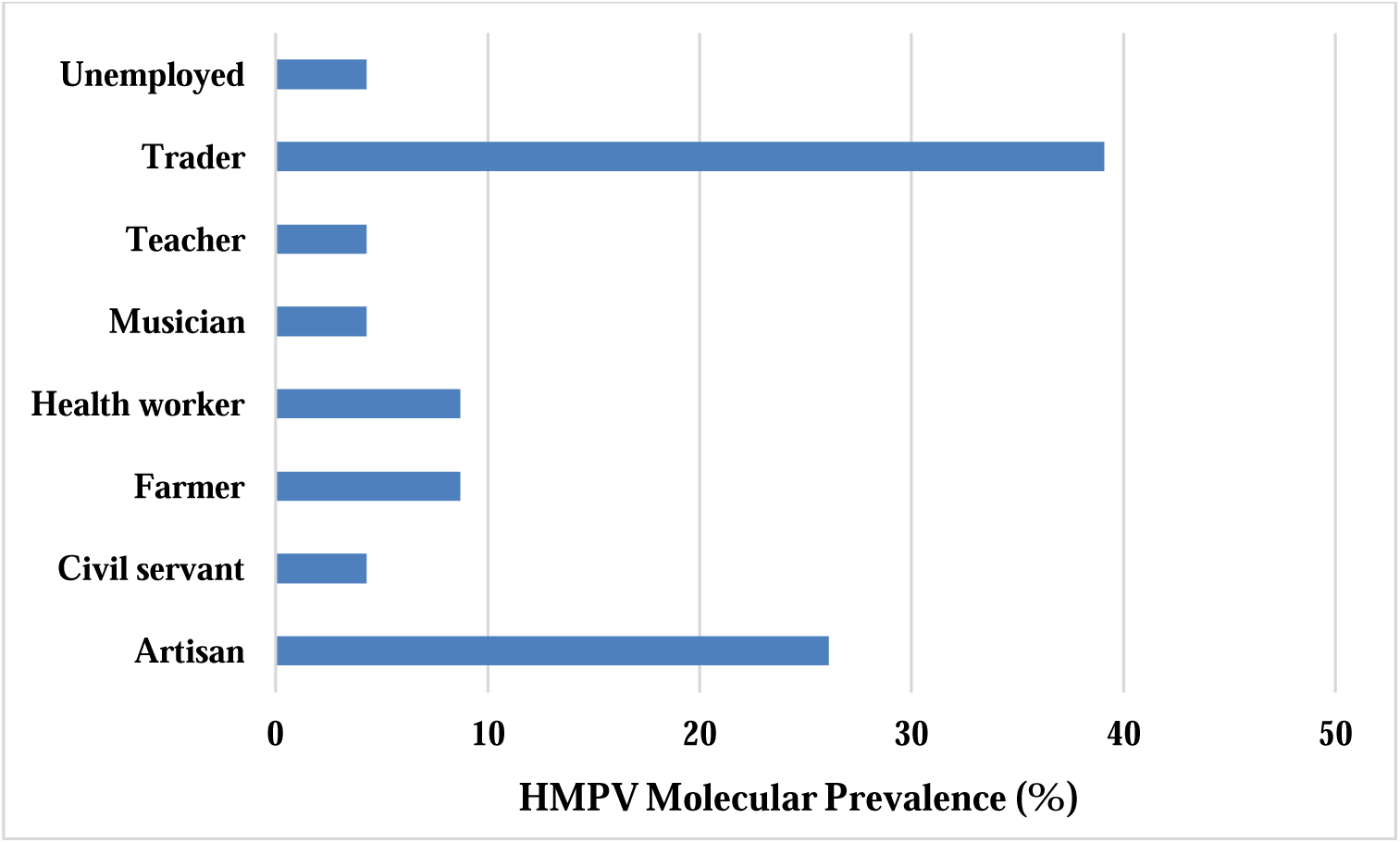
Distribution of HMPV molecular prevalence (%) among the general population in Lagos.

**Table 2:**
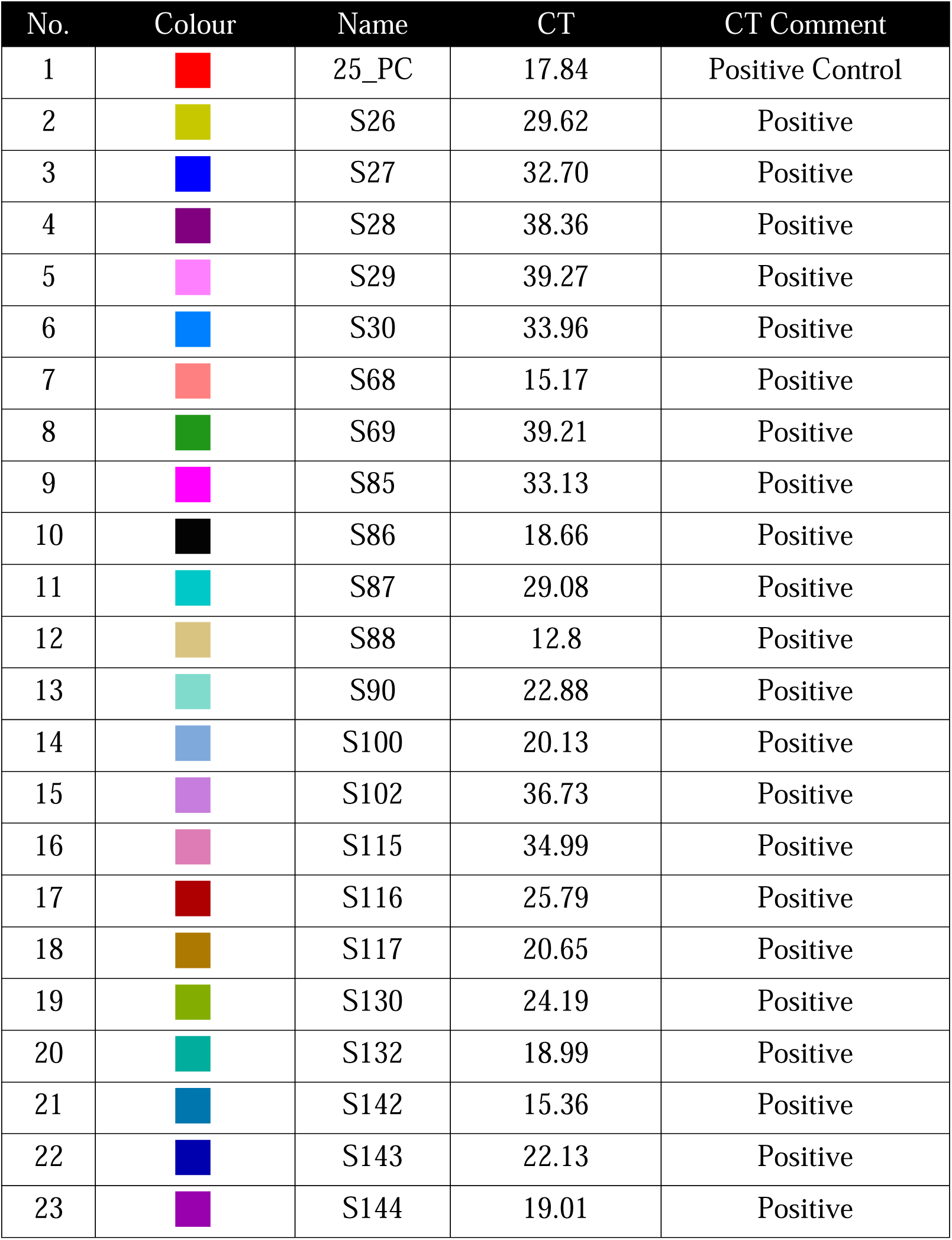
Distribution of CT-values of HMPV positive patients.

| No. | Colour | Name | CT | CT Comment |
| --- | --- | --- | --- | --- |
| 1 |  | 25_PC | 17.84 | Positive Control |
| 2 |  | S26 | 29.62 | Positive |
| 3 |  | S27 | 32.70 | Positive |
| 4 |  | S28 | 38.36 | Positive |
| 5 |  | S29 | 39.27 | Positive |
| 6 |  | S30 | 33.96 | Positive |
| 7 |  | S68 | 15.17 | Positive |
| 8 |  | S69 | 39.21 | Positive |
| 9 |  | S85 | 33.13 | Positive |
| 10 |  | S86 | 18.66 | Positive |
| 11 |  | S87 | 29.08 | Positive |
| 12 |  | S88 | 12.8 | Positive |
| 13 |  | S90 | 22.88 | Positive |
| 14 |  | S100 | 20.13 | Positive |
| 15 |  | S102 | 36.73 | Positive |
| 16 |  | S115 | 34.99 | Positive |
| 17 |  | S116 | 25.79 | Positive |
| 18 |  | S117 | 20.65 | Positive |
| 19 |  | S130 | 24.19 | Positive |
| 20 |  | S132 | 18.99 | Positive |
| 21 |  | S142 | 15.36 | Positive |
| 22 |  | S143 | 22.13 | Positive |
| 23 |  | S144 | 19.01 | Positive |

## 4. DISCUSSION

The cross-sectional study conducted in Lagos provides an important insight into the prevalence and distribution of Human Metapneumovirus (HMPV) among individuals with underlying conditions. The findings of this research contribute tremendously to our previous understanding of HMPV epidemiology and also have important implications for public health strategies.

A major finding of this study is the overall HMPV molecular prevalence of 15.3% in the general population. This prevalence rate is similar to the study by Arshad *et al*. in Pakistan, where 16.5% of 127 children with respiratory symptoms tested positive for HMPV. Also, the study by Takayama *et al*. in Japan reported a similar prevalence of 16.05% among 380 patients with respiratory illnesses. Many studies have also recorded high prevalence (Souza, 2012, El Chaer, 2017, Yan, 2017, Taniguchi, 2019, Zhao, 2020). This consistency in prevalence rates across geographically diverse populations indicates that HMPV is a globally significant pathogen with a steady presence in different regions.

In contrary, the prevalence reported in this study is higher than that reported elsewhere. For instance, Hindupur, 2022, and Shuaibu, 2018 found a prevalence of only 4% and 4.8% in children under 5 years old in India and Nigeria respectively. Likewise, Akinloye *et al*. (2011) reported a prevalence of 3.7% among children under five years in Lagos. A prevalence of 5.1% was also reported in Germany (Huck et al, 2006) and 5% in Vietnam (Yoshida et al, 2008). The lower prevalence in these studies could be attributed to the younger age groups involved, as the current study suggests that HMPV prevalence increases with age, particularly among individuals over 55 years. This significant variation in prevalence rates across different geographical locations emphasizes the need for local epidemiological studies in understanding the burden of HMPV in specific populations.

There is a significant age-related outcome in HMPV prevalence in this study, with the highest prevalence observed in the age group above 55 years (34.8%). This finding is however similar to the study by Wei *et al*. in Kuala Lumpur, where the majority of HMPV-positive cases were adults and elderly individuals. A study by Loubet in 2021 in France also recorded prevalence among adults of over 60 years who had chronic conditions, frequent respiratory and cardiac chronic diseases, and frequently presented complications. This finding also agrees with the studies of Falsey, 2003, Hamelin, 2005, Walsh, 2008, Haas, 2012, and Hasvold, 2016 where high prevalence was recorded in older adults. The increase in prevalence in older populations could be due to a combination of factors, including a weakened immune system and higher exposure to environments where respiratory infections are prevalent, such as healthcare settings. This finding also differs from many previous studies that targeted HMPV in pediatric populations. For example, studies in Taiwan by Wei, 2012, Bangladesh by Rahman, 2018 and many numerous studies have reported significant HMPV positivity rates among children (Xepapadaki, 2004, Greensil, 2003, Richard, 2008, Semple, 2005, Foulongne, 2006, König, 2004, García-García, 2006, Wolf, 2006, Yan, 2016, Taniguchi, 2019). This shows the need for increased attention to HMPV in pediatric and geriatric care and indicates the importance of considering HMPV in differential diagnoses for respiratory infections in children and older adults.

The male predominance reported in the Lagos study (56.6%) agrees with other research, such as the study by Yi *et al*. in China, where a higher number of HMPV cases were reported among males (65.8%). Scheuerman in 2016 recorded a male prevalence of 67% and El Chaer reported a male prevalence of 60% in 2017. The male predominance could be due to various socio-cultural factors, including occupational exposure, as seen in this study where traders, who are often male, have the highest prevalence. However this is in contrary to the findings of Wei *et al* where females 61.6% had the highest prevalence of HMPV. Studies by Paranhos-Baccalà, 2008, Lenahan, 2017 Emont, 2019, and Schwartz, 2020 also reported high prevalence in females. Interestingly, a study by Celebi in 2021 reported equal number of prevalence (6 each) from both males and females.

In this study, the high prevalence of HMPV among patients with co-infections or comorbidity is worrisome. The most significant co-infection was with malaria, where 43.3% of the HMPV-positive cases also had malaria. This is a novel finding, as the interaction between HMPV and malaria has not been extensively documented in literature. Additionally, other co-morbid conditions such as high blood pressure (26.1%), diabetes (21.7%), tuberculosis (17.4%), and pneumonia (13.3%) were also significant.

These reports align with global studies indicating a high prevalence of co-infections in HMPV-positive patients, although the types of co-infections vary across regions. For example, the study by Moe et al. in Norway reported, 47.3% of HMPV positive patients also had respiratory syncytial virus (RSV), and 0.6% had both HMPV and RSV, indicating a significant overlap between these two respiratory viruses. This is similar to this study, where respiratory conditions such as pneumonia were common among HMPV-positive patients; this suggests that HMPV might often co-occur with other respiratory pathogens. Interestingly, no viral co-infections between members of the orthomyxoviruses (influenza) and the paramyxoviruses (parainfluenza, RSV, HMPV) were seen in a study by Franz, 2010.

Also, the study by Arnott *et al*. in Cambodia reported that a significant number of patients presented with hyperleukocytosis, suggestive of a potential bacterial co-infection. This pattern of viral co-infections agrees with the findings in this study, where patients with underlying conditions such as tuberculosis and pneumonia were found to have a high prevalence of HMPV. The presence of multiple pathogens could exacerbate respiratory symptoms, leading to more severe clinical outcomes. Association of HMPV with co infections was also reported by Yoshida *et al*. in Vietnam and Wei *et al* in Malaysia, as well as by Williams, 2005, Egli, 2012, Renaud, 2013, Souza, 2013, Scheuerman, 2016, Yan, 2016, El Chaer, 2017, Loubet, 2020, Jethani, 2021.

Our findings of high HMPV prevalence among patients with co-infections and comorbidities are consistent with global trends. The interaction between HMPV and other pathogens, such as malaria and respiratory viruses suggests that HMPV may exacerbate existing health conditions or that individuals with these conditions are more prone to HMPV infection. This highlights the need for comprehensive diagnostic approaches in clinical settings to identify and manage multiple infections effectively.

The study also reveals significant geographical variation in HMPV prevalence across different regions in Lagos State with the highest prevalence recorded in Badagry (39.1%) and the lowest in Alimosho (26.1%). This variation could be influenced by factors such as population density, healthcare access, and environmental conditions, which are known to affect the transmission pattern of respiratory viruses. For instance, Badagry, being a border town has higher human traffic, which could contribute to the increased prevalence of HMPV.

Occupationally, this study reports the highest prevalence of HMPV (39.1%) among traders, which could be due to their increased interaction with the public and possible exposure to the virus in crowded markets. The low prevalence among civil servants, musicians, teachers, and the unemployed (4.3%) suggests that these groups might have lower exposure risks, possibly due to their work environments or socio-economic factors that limit their contact with large groups of people.

The high prevalence of HMPV reported in this study is closer to the reports in regions with similar socio-economic conditions and healthcare challenges, such as the study in Islamabad, Pakistan. In contrast, studies conducted in more developed regions or among populations with better healthcare access, such as Wei *et al*.’s study in Taiwan, generally reported lower prevalence rates. This suggests that socio-economic and healthcare-related factors play a significant role in the epidemiology of HMPV. However, part of the limitations of our current findings is the inability to characterize the circulating genotypes and establish the seasonality of HMPV in this part of the world. We therefore strongly recommend the genotyping of circulating strains and the need to understand the mechanisms of comorbidities with important aetiologies like *Plasmodium spp*. and *Mycobacterium tuberculosis,* as well as underlying conditions of hypertension and diabetes.

### Conclusion

This study provides important insights into the molecular detection and epidemiology of HMPV in Lagos, Nigeria. The findings highlight the significant prevalence of HMPV, particularly among older adults and individuals with comorbid conditions. The overall 15.3% molecular detection of HMPV among children and adults from different communities in Lagos accentuates the importance of HMPV as a viral pathogen causing respiratory infections in the metropolis. Hence, the need for a continuous surveillance and genomic analysis of this viral aetiology especially among individuals with underlying diseases.

The geographical and occupational variations in HMPV prevalence observed in this study propose the need for targeted public health interventions to curb the spread of HMPV, especially in high-risk groups. When compared with existing literature, the prevalence of HMPV in Lagos is relatively high, reflecting the unique socio-economic and healthcare dynamics of the region. Further research is needed to explore the relationship between HMPV and co-infections like malaria, as well as the impact of socio-demographic factors on HMPV transmission dynamics and outcomes. The findings highlight the need for increased awareness of HMPV among healthcare providers, especially when treating patients with co-morbidities such as malaria, hypertension, and diabetes. Future research should focus on longitudinal studies to understand the seasonal patterns of HMPV in the region, as well as investigations into the clinical outcomes and potential complications of HMPV infection in different risk groups in broader population.

The results of this study have important implications for public health policy and clinical practice. They suggest the need for further surveillance systems for HMPV, particularly in regions with high prevalence. Inclusion of HMPV testing in routine diagnostic panels for respiratory infections, especially in high-risk groups, development of targeted prevention strategies for vulnerable populations, such as the elderly and those with specific co-morbidities and further research into potential vaccines or antiviral treatments for HMPV, given its significant prevalence and impact on vulnerable populations.

## Data Availability

All data produced in the present study are available upon reasonable request to the authors.

## Acknowledgements

The authors would like to acknowledge the Lagos State University management especially the Department of Microbiology and all the team members in the influenza and other respiratory tract virus (IORTV) Research, and the University teaching hospital ethical review board for their various support and for granting the approval to carry out this research. All the different participants are specifically acknowledged for agreeing to be part of this current research.

## Ethics Statement

The study ethics permission was sought after from the Heads of Health Centers where the samples were obtained while the ethical approval was obtained from LASUTH (Lagos State University Teaching Hospital) with the registration number NHREC04/04/2008 and reference number LREC/06/10/1272, respectively.

## Author contributions

AAA carried out conception, design of the work, acquisition, analysis, interpretation of data; drafted the work and revised it.

HOO participated in acquisition, carried out analysis of the software used in the work, interpretation of data, participated in the initial draft of the work and substantively revised it.

SAS, analysis, interpretation of data, and final draft.

TDA participated in acquisition, analysis, interpretation of data, and initial draft. GTD participated in acquisition, analysis, interpretation of data, and initial draft. KOA participated in acquisition, interpretation of data and quality control.

All authors approved the submitted version.

## Data Availability Statement

The datasets used and/or analyzed to support the findings of this study are available upon reasonable request from the corresponding author.

## Informed Consent

Oral and written informed consents were obtained from all the participants and appropriately documented.

## References

1. Agapov, E., Sumino, K.C., Gaudreault-Keener, M., Storch, G.A. and Holtzman, M.J., 2006. Genetic variability of human metapneumovirus infection: evidence of a shift in viral genotype without a change in illness. Journal of Infection, 193, p.396.

2. Akinloye, O.M., Rönkkö, E., Savolainen-Kopra, C., Ziegler, T., Iwalokun, B.A., Deji-Agboola, M.A., Oluwadun, A., Roivainen, M., Adu, F.D. and Hovi, T., 2011. Specific viruses detected in Nigerian children in association with acute respiratory disease. Journal of Tropical Medicine, 2011, p.690286. doi: 10.1155/2011/690286.

3. Alvarez, R. and Tripp, R.A., 2005. The immune response to human metapneumovirus is associated with aberrant immunity and impaired virus clearance in BALB/c mice. Journal of Virology, 79, pp.5971–5978.

4. Arnott, A., Vong, S., Sek, M., Naughtin, M., Beauté, J., Rith, S., Guillard, B., Deubel, V. and Buchy, P., 2013. Genetic variability of human metapneumovirus amongst an all ages population in Cambodia between 2007 and 2009. Infection, Genetics and Evolution, 15, pp.43–52. doi: 10.1016/j.meegid.2011.01.016.

5. Arshad, Y., Rana, M.S., Ikram, A., Salman, M., Aamir, U.B., Zaidi, S.S.Z., Alam, M.M., Sharif, S., Shaukat, S., Khurshid, A., Hakim, R., Mujtaba, G., Umair, M. and Bostan, S.S.N., 2022. Molecular detection and genetic characterization of human metapneumovirus strains circulating in Islamabad, Pakistan. Scientific Reports, 12, p.2790. doi: 10.1038/s41598-022-06537-5.

6. Boivin, G., Abed, Y., Pelletier, G. et al., 2002. Virological features and clinical manifestations associated with human metapneumovirus: a new paramyxovirus responsible for acute respiratory tract infections in all age groups. Journal of Infectious Diseases, 186(9), pp.1330–1334.

7. Boivin, G., De Serres, G., Côté, S., Gilca, R., Abed, Y., Rochette, L., Bergeron, M.G. and Déry, P., 2003. Human metapneumovirus infections in hospitalized children. Emerging Infectious Diseases, 9(6), pp.634–640. doi: 10.3201/eid0906.030017.

8. Bosis, S., Esposito, S., Niesters, H.G., Crovari, P., Osterhaus, A.D. and Principi, N., 2005. Impact of human metapneumovirus in childhood: comparison with respiratory syncytial virus and influenza viruses. Journal of Medical Virology, 75(1), pp.101–104. doi: 10.1002/jmv.20243.

9. Caracciolo, S., Minini, C., Colombrita, D., Rossi, D., Miglietti, N., Vettore, E., Caruso, A. and Fiorentini, S., 2008. Human metapneumovirus infection in young children hospitalized with acute respiratory tract disease: virologic and clinical features. Pediatric Infectious Disease Journal, 27(5), pp.406–412. doi: 10.1097/INF.0b013e318162a164.

10. Carneiro, B.M., Yokosawa, J., Arbiza, J., Costa, L.F., Mirazo, S., Nepomuceno, L.L., Oliviera, T.F., Goulart, L.R., Vieira, C.U., Freitas, G.R., Paula, N.T. and Quieroz, D.A., 2009. Detection of all four human metapneumovirus subtypes in nasopharyngeal specimens from children with respiratory disease in Uberlândia, Brazil. Journal of Medical Virology, 81, pp.1814–1818.

11. Çelebi, Ö. and Çelebi, D., 2021. Viral respiratory tract pathogens during the COVID-19 pandemic. Eurasian Journal of Medicine, 53(2), pp.123–126. doi: 10.5152/eurasianjmed.2021.20459.

12. Cespedes, P.F., Gonzalez, P.A. and Kalergis, A.M., 2013. Human metapneumovirus keeps dendritic cells from priming antigen-specific naïve T-cells. Immunology, 139, pp.366–376.

13. Chan, P.C., Wang, C.Y., Wu, P.S., Chang, P.Y., Yang, T.T. and Chiang, Y.P., 2007. Detection of human metapneumovirus in hospitalized children with acute respiratory tract infection using real-time RT-PCR in a hospital in northern Taiwan. Journal of the Formosan Medical Association, 106, pp.16–24.

14. Chow, W.Z., Chan, Y.F., Oong, X.Y., Ng, L.J., Nor’E, S.S., Ng, K.T., Chan, K.G., Hanafi, N.S., Pang, Y.K., Kamarulzaman, A. and Tee, K.K., 2016. Genetic diversity, seasonality and transmission network of human metapneumovirus: identification of a unique sub-lineage of the fusion and attachment genes. Scientific Reports, 6, p.27730. doi: 10.1038/srep27730.

15. Chung, J.Y., Han, T.H., Kim, S.W. and Hwang, E.S., 2008. Genotype variability of human metapneumovirus, South Korea. Journal of Medical Virology, 80, pp.902–905.

16. Corti, D., Bianchi, S., Vanzetta, F., Minola, A., Perez, L. and Agatic, G., 2013. Cross-neutralization of four paramyxoviruses by a human monoclonal antibody. Nature, 501, pp.439–443.

17. Domachowske, J., 2018. Pediatric human metapneumovirus. Medscape. Available at: https://emedicine.medscape.com/article/972492-overview [Accessed 16 March 2020].

18. Duchamp, M.B., Lina, B., Trompette, A., Moret, H., Motte, J. and Andreoletti, L., 2005. Detection of human metapneumovirus RNA sequence in nasopharyngeal aspirates of young French children with acute bronchiolitis by real-time reverse transcriptase PCR and phylogenetic analysis. Journal of Clinical Microbiology, 43, pp.1411–1414.

19. Egli, A., Bucher, C., Dumoulin, A., Stern, M., Buser, A., Bubendorf, L. et al., 2012. Human metapneumovirus infection after allogeneic hematopoietic stem cell transplantation. Infection, 40(6), pp.677–684.

20. El Chaer, F., Shah, D.P., Kmeid, J., Ariza-Heredia, E.J., Hosing, C.M., Mulanovich, V.E. and Chemaly, R.F., 2017. Burden of human metapneumovirus infections in patients with cancer: risk factors and outcomes. Cancer, 123(12), pp.2329–2337. doi: 10.1002/cncr.30599.

21. Emont, J., Chung, K. and Rouse, D., 2019. A report of two cases of human metapneumovirus infection in pregnancy involving superimposed bacterial pneumonia and severe respiratory illness. Journal of Clinical Gynecology and Obstetrics, North America, December. Available at: https://jcgo.org/index.php/jcgo/article/view/573/387 [Accessed 7 September 2024].

22. Falsey, A.R. and Walsh, E.E., 2006. Viral pneumonia in older adults. Clinical Infectious Diseases, 42(4), pp.518–524.

23. Falsey, A.R., Erdman, D., Anderson, L.J. and Walsh, E.E., 2003. Human metapneumovirus infections in young and elderly adults. Journal of Infectious Diseases, 187, pp.785–790.

24. Feltes, T.F., Cabalka, A.K., Meissner, H.C., Piazza, F.M., Carlin, D.A., Top, F.H. Jr, Connor, E.M. and Sondheimker, H.M., 2003. Palivizumab prophylaxis reduces hospitalization due to respiratory syncytial virus in young children with hemodynamically significant congenital heart disease. Journal of Pediatrics, 143, pp.532–540.

25. Foulongne, V., Guyon, G., Rodière, M. and Segondy, M., 2006. Human metapneumovirus infection in young children hospitalized with respiratory tract disease. Pediatric Infectious Disease Journal, 25, pp.354–359.

26. Franz, A., Adams, O., Willems, R., Bonzel, L., Neuhausen, N., Schweizer-Krantz, S., Ruggeberg, J.U., Willers, R., Henrich, B., Schroten, H. and Tenenbaum, T., 2010. Correlation of viral load of respiratory pathogens and co-infections with disease severity in children hospitalized for lower respiratory tract infection. Journal of Clinical Virology, 48(4), pp.239–245. doi: 10.1016/j.jcv.2010.05.007.

27. Freymuth, F., Vabret, A., Gouarin, S., Petitjean, J., Charbonneau, P., Lehoux, P., Galateau-Salle, F., Tremolieres, F., Carette, M.F., Mayaud, C. and Mosnier, A., 2004. Epidemiology and diagnosis of respiratory syncytial virus in adults. [Journal name not provided], 21(1), pp.35–42.

28. Freymuth, F., Vabret, A., Legrand, L., Etaradossi, N., Lafay-Delaire, F., Brouard, J. and Guillois, B., 2003. Presence of the new human metapneumovirus in French children with bronchiolitis. Pediatric Infectious Disease Journal, 22, pp.92–94.

29. Galiano, M., Trento, A., Ver, L., Carballal, G. and Videla, C., 2006. Genetic heterogeneity of G and F protein gene from Argentinean human metapneumovirus strain. Journal of Medical Virology, 78, pp.631–637.

30. García-García, M.L., Calvo, C., Pérez-Breña, P., De Cea, J.M., Acosta, B. and Casas, I., 2006. Prevalence and clinical characteristics of human metapneumovirus infections in hospitalized infants in Spain. Pediatric Pulmonology, 41, pp.863–871.

31. Garrett, A., Chu, H.Y., et al., 2021. Human metapneumovirus infection and genotyping of infants in rural Nepal. Journal of the Pediatric Infectious Diseases Society, 10(4), pp.408–416.

32. Graci, J.D. and Cameron, C.E., 2006. Mechanism of action of ribavirin against distinct viruses. Reviews in Medical Virology, 16, pp.37–48.

33. Greensill, J., McNamara, P.S., Dove, W., Flanagan, B., Smyth, R.L. and Hart, C.A., 2003. Human metapneumovirus in severe respiratory syncytial virus bronchiolitis. Emerging Infectious Diseases, 9, pp.372–375.

34. Haas, L.E.M., de Rijk, N.X. and Thijsen, S.F.T., 2012. Human metapneumovirus infections on the ICU: a report of three cases. Annals of Intensive Care, 2, p.30

35. Hamelin, M.E., Cotu, S., Laforge, J., Lampron, N., Bourbeau, J. and Weiss, K. et al., 2005. Human metapneumovirus infection in adults with community-acquired pneumonia and exacerbation of chronic obstructive pulmonary disease. Clinical Infectious Diseases, 41, pp.498–502.

36. Hamelin, M.E., Couture, C., Saxkett, M.K. and Biovin, G., 2007. Enhanced lung disease and Th2 response following human metapneumovirus infection in mice immunized with the inactivated virus. Journal of General Virology, 88, pp.3391–3400.

37. Hasvold, J., Sjoding, M., Pohl, K., Cooke, C.R. and Hyzy, R.C., 2016. The role of human metapneumovirus in the critically ill adult patient. Journal of Critical Care, 31, pp.233–237.

38. Herd, K.A., Mahalingam, S., Mackay, I.M., Nissen, M., Sloots, T.P. and Tindle, R.W., 2006. Cytotoxic T-lymphocyte epitope vaccination protects against human metapneumovirus infection and disease in mice. Journal of Virology, 80, pp.2034–2044.

39. Herfst, S.M., Schrauwen, E.J., Graff, M., Van Amerongen, G., Van den Hoogen, B.G. and Swart, R.L., 2008. Immunogenicity and efficacy of two candidate human metapneumovirus vaccines in cynomolgus macaques. Vaccine, 26, pp.4224–4230.

40. Hindupur, A., Menon, T. and Dhandapani, P., 2022. Molecular investigation of human metapneumovirus in children with acute respiratory infections in Chennai, South India, from 2016–2018. Brazilian Journal of Microbiology, 53(2), pp.655–661. doi: 10.1007/s42770-022-00689-2.

41. Huck, B., Scharf, G., Neumann-Haefelin, D., Puppe, W., Weigl, J. and Falcone, V., 2006. Novel human metapneumovirus sublineage. Emerging Infectious Diseases, 12(1), pp.147–150. doi: 10.3201/eid1201.050772.

42. Jallow, M.M., Fall, A., Kiori, D., Sy, S., Goudiaby, D., Barry, M.A., Fall, M., Ndiaye Niang, M. and Dia, N., 2019. Epidemiological, clinical and genotypic features of human metapneumovirus in patients with influenza-like illness in Senegal. BMC Infectious Diseases, 19, p.457.

43. Jethani, J., Samad, S., Kumar, P., Angel, B., Wig, N., Choudhary, A., Brijwal, M., Kumar, L. and Dar, L., 2021. Human metapneumovirus infection in haematopoietic stem cell transplant recipients: a case series. Virusdisease, 32(1), pp.140–145. doi: 10.1007/s13337-021-00670-x.

44. Kahn, J.S., 2006. Epidemiology of human metapneumovirus. Clinical Microbiology Reviews, 19, pp.546–557.

45. Kato, T., Mizokami, M. and Mukaide, M., 2000. [Title not provided]. Journal of Clinical Microbiology, 38, pp.94–98.

46. Kolawole, O., Oguntoye, M., Dam, T. and Chunara, R., 2013. Etiology of respiratory tract infections in the community and clinic in Ilorin, Nigeria. BMC Research Notes, 10, p.712.

47. König, B., König, W., Arnold, R., Werchau, H., Ihorst, G. and Forster, J., 2004. Prospective study of human metapneumovirus infection in children less than 3 years of age. Journal of Clinical Microbiology, 42, pp.4632–4635.

48. Koo, H.J., Lim, S., Choe, J., Choi, S.H., Sung, H. and Do, K.H., 2018. Radiographic and CT features of viral pneumonia. Radiographics, 38(3), pp.719–739.

49. Lara, M., Ghosh, A. and Plata, A., 2012. Critical role of MDA5 in interferon response induced by human metapneumovirus infection in dendritic cells and in vivo. Journal of Virology, 87, pp.1242–1251.

50. Lee LG, Connell CR, Bloch W. Allelic discrimination by nick-translation PCR with fluorogenic probes. Nucleic Acids Res. 1993 Aug 11;21(16):3761–6. doi: 10.1093/nar/21.16.3761. PMID: 8367293; PMCID: PMC309885.

51. Lenahan, J.L., Englund, J.A., Katz, J., Kuypers, J., Wald, A., Magaret, A., Tielsch, J.M., Khatry, S.K., LeClerq, S.C., Shrestha, L., Steinhoff, M.C. and Chu, H.Y., 2017. Human metapneumovirus and other respiratory viral infections during pregnancy and birth, Nepal. Emerging Infectious Diseases, 23(8), pp.1341–1349. doi: 10.3201/eid2308.161358.

52. Levy, C., Aerts, L., Hamelin, M.E., Granier, C.M., Szcesi, J. and Lavillette, D., 2013. Virus-like particle vaccine induces cross protection against human metapneumovirus infection in mice. Vaccine, 31, pp.2778–2785.

53. Liu, P., Shu, Z., Qin, X., Duo, Y., Zhao, Y. and Zhao, X., 2013. A live attenuated human metapneumovirus vaccine strain provides complete protection against homologous viral infection and cross-protection against heterologous viral infection in BALB/c mice. Clinical Vaccine Immunology, 20, pp.1246–1254.

54. Loubet, P., Mathieu, P., Lenzi, N., Galtier, F., Lainé, F., Lesieur, Z., Vanhems, P., Duval, X., Postil, D., Amour, S., Rogez, S., Lagathu, G., L’Honneur, A.S., Foulongne, V., Houhou, N., Lina, B., Carrat, F. and Launay, O.; Fluvac Study Group, 2021. Characteristics of human metapneumovirus infection in adults hospitalized for community-acquired influenza-like illness in France, 2012–2018: a retrospective observational study. Clinical Microbiology and Infection, 27(1), pp.127.e1–127.e6. doi: 10.1016/j.cmi.2020.04.005.

55. Matsuzaki, Y., Itagaki, T., Ikeda, T., Aoki, Y., Abiko, C. and Mizuta, K., 2012. Human metapneumovirus among family members. [Journal not provided], pp.1–6.

56. Mizuta, K., Abiko, C., Akoi, Y., Ikeda, T., Matsuza, Y. and Itagaki, T. et al., 2013. Seasonal patterns of respiratory syncytial virus, influenza A virus, human metapneumovirus and parainfluenza virus type 3 on the basis of virus isolation data between 2004 and 2011 in Yamagata, Japan. Japanese Journal of Infectious Diseases, 66, pp.140–145.

57. Moe, N., Krokstad, S., Stenseng, I.H., Christensen, A., Skanke, L.H., Risnes, K.R., Nordbø, S.A. and Døllner, H., 2017. Comparing human metapneumovirus and respiratory syncytial virus: viral co-detections, genotypes and risk factors for severe disease. PLoS One, 12(1), p.e0170200. doi: 10.1371/journal.pone.0170200.

58. Mona, S., Mohamed, E., Reiche, J., Jacobsen, S.M., Thanit, A.G. and Badary, M.S., 2014. Molecular analysis of human metapneumovirus detected in patients with lower respiratory tract infection in Upper Egypt. International Journal of Microbiology, 2014, p.290793.

59. Monto, A.S., 2002. Epidemiology of viral respiratory infections. American Journal of Medicine, 112, pp.4S–12S.

60. Mullins, J.A., Erdman, D.D., Weinberg, G.A. et al., 2004. Human metapneumovirus infection among children hospitalized with acute respiratory illness. Emerging Infectious Diseases, 10(4), pp.700–705.

61. Nidaira, M., Taira, K., Hamabata, H., Kawaki, T., Gushi, K., Mahoe, Y., Maeshiro, N., Azama, Y., Okano, S., Kyan, H., Kudaka, J., Tsukagoshi, H., Noda, M. and Kimura, H., 2012. Molecular epidemiology of human metapneumovirus from 2009 to 2011 in Okinawa, Japan. Japanese Journal of Infectious Diseases, 65(4), pp.337–340. doi: 10.7883/yoken.65.337.

62. Osterhaus, A. and Fouchier, R., 2003. Human metapneumovirus in the community. The Lancet, 361, pp.890–891.

63. Owor, B.E., Ndegwa, G., Chebet, L., Wammanda, R., Nyagah, C. and Nokes, J., 2016. Human metapneumovirus epidemiological and evolutionary patterns in coastal Kenya. BMC Infectious Diseases, 16, p.301.

64. Paranhos-Baccalà, G., Komurian-Pradel, F., Richard, N., Vernet, G., Lina, B. and Floret, D., 2008. Mixed respiratory virus infections. Journal of Clinical Virology, 43(4), pp.407–410. doi: 10.1016/j.jcv.2008.08.010.

65. Peiris, J.S., Tang, W.H., Chan, K.H., Khong, P.L., Guan, Y., Lau, Y.L. and Chiu, S.S., 2003. Children with respiratory disease associated with metapneumovirus in Hong Kong. Emerging Infectious Diseases, 9, pp.628–633.

66. Peret, T.C., Boivin, G., Li, Y., Couillard, M., Humphrey, C., de Osterhaus, A.D. and Erdman, D.D., 2002. Characterization of human metapneumoviruses isolated from patients in North America. Journal of Infectious Diseases, 185, pp.1660–1663.

67. Pilger, D.A., Cantarelli, V.V., Amentea, S.L. and Leistner-Segals, [initials not provided], 2011. Detection of human bocavirus and human metapneumovirus by real-time PCR from patients with respiratory symptoms in Southern Brazil. Memórias do Instituto Oswaldo Cruz, 106, pp.56–60.

68. Rahman MZ, Sumiya M, Sahabuddin M, Pell LG, Gubbay JB, Rahman R, Momtaz F, Azmuda N, Shanta SS, Jahan I, Rahman M, Mahmud AA, Roth DE, Morris SK. Genetic characterization of human metapneumovirus identified through community and facility-based surveillance of infants in Dhaka, Bangladesh. J Med Virol. 2019 Apr;91(4):549–554. doi: 10.1002/jmv.25351. Epub 2018 Nov 13. PMID: 30372530.

69. Reicher, J., Jacobsen, D., Neubauer, K., Hafemann, S., Nishe, A., Milde, J., Wplff, T. and Schweiger, B., 2014. Human metapneumovirus: insights from a ten-year molecular and epidemiological analysis in Germany. PLOS One, 9, p.e88324.

70. Renaud, C., Xie, H., Seo, S., Kuypers, J., Cent, A., Corey, L. et al., 2013. Mortality rates of human metapneumovirus and respiratory syncytial virus lower respiratory tract infections in hematopoietic cell transplantation recipients. Biology of Blood and Marrow Transplantation, 19(8), pp.1220–1226.

71. Richard, N., Komurian-Pradel, F., Javouhey, E., Perret, M., Rajoharison, A. and Bagnaud, A., 2008. The impact of dual viral infection in infants admitted to a pediatric intensive care unit associated with severe bronchiolitis. Pediatric Infectious Disease Journal, 27(3), pp.213–217.

72. Russ, B.E., Olshanksy, M., Smallwood, H.S., Li, J., Denton, A.E., Prier, J.E., Stock, A.T., Croom, H.A., Cullen, J.G., Nguyen, M.L., Rowe, S., Olson, M.R., Finkelstein, D.B., Kelso, A., Thomas, P.G., Speed, T.P., Rao, S. and Turner, S.J., 2014. Distinct epigenetic signatures delineate transcriptional programs during virus-specific CD8(+) T cell differentiation. Immunity, 41(5), pp.853–865.

73. Scheuerman, O., Barkai, G., Mandelboim, M., Mishali, H., Chodick, G. and Levy, I., 2016. Human metapneumovirus (hMPV) infection in immunocompromised children. Journal of Clinical Virology, 83, pp.12–16. doi: 10.1016/j.jcv.2016.06.006.

74. Schickli, J.H., Kaur, J., Machphaik, M., Guazzetta, J.M., Spaete, R.R. and Tang, R.S., 2008. Deletion of human metapneumovirus M2-2 increases mutation frequency and attenuates growth in hamsters. Virology Journal, 5, p.69.

75. Schwartz, D.A. and Dhaliwal, A., 2020. Infections in pregnancy with COVID-19 and other respiratory RNA virus diseases are rarely, if ever, transmitted to the fetus: experiences with coronaviruses, HPIV, hMPV, RSV, and influenza. Archives of Pathology & Laboratory Medicine, [ahead of print]. doi: 10.5858/arpa.2020-0211-SA.

76. Semple, M.G., Cowell, A., Dove, W., Greensill, J., McNamara, P.S. and Halfhide, C., 2005. Dual infection of infants by human metapneumovirus and respiratory syncytial virus is strongly associated with severe bronchiolitis. Journal of Infectious Diseases, 191, pp.382–386.

77. Shafagati, N. and Williams, J., 2018. Human metapneumovirus – what we know now. F1000Research, 7, p.135. doi: 10.12688/f1000research.12625.1.

78. Shuaibu, A.H. and Hamal, N.S., 2018. Molecular and conventional methods of detecting respiratory viruses among children with lower respiratory tract infection in Sokoto, Nigeria. Bo Medical Journal, 15(1), pp.65–76.

79. Somayeh, S., Jila, Y., Nazanin, Z., Talat, M. and Mohammed, F., 2020. Prevalence of human metapneumovirus infections in Iran: a systematic review and meta-analysis. Fetal and Pediatric Pathology, 40, pp.663–673.

80. Souza, J.S., Watanabe, A., Carraro, E., Granato, C. and Bellei, N., 2013. Severe metapneumovirus infections among immunocompetent and immunocompromised patients admitted to hospital with respiratory infection. Journal of Medical Virology, 85(3), pp.530–536. doi: 10.1002/jmv.23477.

81. Takayama, I., Semba, S., Yokono, K., Saito, S., Nakauchi, M., Kubo, H., Kaida, A., Shiomi, M., Terada, A., Murakami, K., Kaji, K., Kiya, K., Sawada, Y., Oba, K., Asai, S., Yonekawa, T., Watanabe, H., Segawa, Y., Notomi, T. and Kageyama, T., 2020. Clinical evaluation of fully automated molecular diagnostic system “Simprova” for influenza virus, respiratory syncytial virus, and human metapneumovirus. Scientific Reports, 10(1), p.13496. [Erratum in: Scientific Reports, 10(1), p.16470. doi: 10.1038/s41598-020-74039-3] doi: 10.1038/s41598-020-70090-2.

82. Tan, X.Q., Zhao, X., Lee, V.J., Loh, J.P., Tan, B.H., Koh, W.H., Ng, S.H., Chen, M.I. and Cook, A.R., 2014. Respiratory viral pathogens among Singapore military servicemen 2009–2012: epidemiology and clinical characteristics. BMC Infectious Diseases, 14, p.204.

83. Tang, R.S., Schickli, J.H., MacPhail, M., Fernandes, F., Bicha, L. and Spaete, J., [year not provided]. Effects of human metapneumovirus and respiratory syncytial virus antigen insertion in two proximal genome positions of bovine/human parainfluenza virus type 3 on virus replication and immunogenicity. Journal of Virology, 77, pp.10819–10828.

84. Taniguchi, A., Kawada, J., Go, K., Fujishiro, N., Hosokawa, Y., Maki, Y., Sugiyama, Y., Suzuki, M., Tsuji, T., Hoshino, S., Muramatsu, H., Kidokoro, H., Kinoshita, F., Hirakawa, A., Takahashi, Y., Sato, Y., Natsume, J. and the Nagoya Collaborative Clinical Research Team, 2019. Comparison of clinical characteristics of human metapneumovirus and respiratory syncytial virus infections in hospitalized young children. Japanese Journal of Infectious Diseases, 72(4), pp.237–242. doi: 10.7883/yoken.JJID.2018.480.

85. Te Wierik, M.J., Nguyen, D.T., Beersma, M.F., Yhijsen, S.F. and Heemstra, K.A., 2012. An outbreak of severe respiratory tract infection caused by human metapneumovirus in a residential care facility for the elderly in Utrecht, the Netherlands. Eurosurveillance, 17, p.20132.

86. Tellier, R., Li, Y., Cowling, B.J. and Tang, J.W., 2019. Recognition of aerosol transmission of infectious agents: a commentary. BMC Infectious Diseases, 19, p.101.

87. Ulbrandt, N.D., Ji, H., Patel, N.K., Barnes, A.S., Wilson, S., Kiener, P.A., Suzich, J. and McCarthy, M.P., 2008. Identification of antibody neutralization epitopes on the fusion protein of human metapneumovirus. Journal of General Virology, 89, pp.3113–3118.

88. Van den Hoogen, B.G., Bestebroer, T.M., Osterhaus, A.D.M.E. and Fouchier, R.A.M., 2002. Analysis of the genomic sequence of a human metapneumovirus. [Journal name not provided], 295, pp.119–132.

89. Van den Hoogen, B.G., Can Doorum, G.J., Fockens, J.C., Cornelissen, J.H., Beyer, W.E., de Groot, R., Osterhaus, A.D. and Fouchier, R.A., 2003. Prevalence and clinical symptoms of human metapneumovirus infection in hospitalized patients. Journal of Infection, 188, pp.1571–1577.

90. Van den Hoogen, B.G., de Jong, J.C., Groen, J., Kuiken, T., de Groot, R., Fouchier, R.A. and Osterhaus, A.D., 2001. A newly discovered human pneumovirus isolated from young children with respiratory tract diseases. Nature Medicine, 7(6), pp.719–724.

91. Van den Hoogen, B.G., Herfst, S., Sprong, L., Cane, P.A., Forleo-Neto, E., de Swart, R.L., Osterhaus, A.D. and Fouchier, R.A., 2004. Antigenic and genetic variability of human metapneumovirus. Emerging Infectious Diseases, 10, pp.658–666.

92. Walsh, E.E., Peterson, D.R. and Falsey, A.R., 2008. Human metapneumovirus infections in adults: another piece of the puzzle. Archives of Internal Medicine, 168, p.2489.

93. Wei, H.Y., Tsao, K.C., Huang, C.G., Huang, Y.C. and Lin, T.Y., 2013. Clinical features of different genotypes/genogroups of human metapneumovirus in hospitalized children. *Journal of Microbiology*, Immunology and Infection, 46(5), pp.352–357. doi: 10.1016/j.jmii.2012.07.007.

94. Williams, J.V., Harris, P.A., Tollefson, S.J., Halburnt-Rush, L.L., Pingsterhaus, J.M., Edwards, K.M., Wright, P.F. and Crowe, J.E. Jr, 2004. Human metapneumovirus and lower respiratory tract disease in otherwise healthy infants and children. New England Journal of Medicine, 350(5), pp.443–450. doi: 10.1056/NEJMoa025472.

95. Williams, J.V., Martino, R., Rabella, N., Otegui, M., Parody, R. and Heck, J.M., 2005. A prospective study comparing human metapneumovirus with other respiratory tract viruses in adults with hematologic malignancies and respiratory tract infections. Journal of Infectious Diseases, 192, pp.1061–1065.

96. Williams, J.V., Wang, C.K., Yang, C.F., Tollefson, S.J., House, F.S., Heck, J.M., Chu, M., Brown, J.B. and Lintao, L.D., 2006. The role of human metapneumovirus in upper respiratory tract infections in children. Journal of Infection, 193, pp.387–395.

97. Wittwer, C.T., Fillmore, G.C. and Garling, D.J., 1990. Minimizing the time required for DNA amplification by efficient heat transfer to small samples. Analytical Biochemistry, [volume not provided], pp.328–331.

98. Wolf, D.G., Greenberg, D., Kalkstein, D., Shemer-Avni, Y., Givon-Lavi, N. and Saleh, N., 2006. Comparison of human metapneumovirus, respiratory syncytial virus and influenza A virus lower respiratory tract infections in hospitalized young children. Pediatric Infectious Disease Journal, 25(4), pp.320–324.

99. Xepapadaki, P., Psarras, S., Bossios, A., Tsolia, M., Gourgiotis, D. and Liapi-Adamidou, G., 2004. Human metapneumovirus as a causative agent of acute bronchiolitis in infants. Journal of Clinical Virology, 30, pp.267–270.

100. Yan, X.L., Li, Y.N., Tang, Y.J., Xie, Z.P., Gao, H.C., Yang, X.M., Li, Y.M., Liu, L.J. and Duan, Z.J., 2017. Clinical characteristics and viral load of respiratory syncytial virus and human metapneumovirus in children hospitalized for acute lower respiratory tract infection. Journal of Medical Virology, 89(4), pp.589–597. doi: 10.1002/jmv.24687.

101. Yi, L., Zou, L., Peng, J., Yu, J., Song, Y., Liang, L., Guo, Q., Kang, M., Ke, C., Song, T., Lu, J. and Wu, J., 2019. Epidemiology, evolution and transmission of human metapneumovirus in Guangzhou, China, 2013–2017. Scientific Reports, 9, p.14022. doi: 10.1038/s41598-019-50340-8.

102. Yoshida, L.M., Suzuki, M., Yamamoto, T., Nguyen, H.A., Nguyen, C.D., Nguyen, A.T., Oishi, K., Vu, T.D., Le, T.H., Le, M.Q., Yanai, H., Kilgore, P.E., Dang, D.A. and Ariyoshi, K., [year not provided]. Viral pathogens associated with acute respiratory infections in central Vietnamese children. Pediatric Infectious Disease Journal, 29(1), pp.75–77. doi: 10.1097/INF.0b013e3181af61e9.

103. Zhao, L.Q., Deng, L., Cao, L., Chen, D.M., Sun, Y., Zhu, R.N., Wang, F., Guo, Q., Zhou, Y.T., Jia, L.P., Huang, H., Kang, X.H., Jin, F.H., Yuan, Y., Zhang, N., De, R. and Qian, Y., 2020. Investigation of pathogenic agents causing acute respiratory tract infections in pediatric patients in a children’s hospital assigned for case screening in Beijing during the outbreak of COVID-19. Zhonghua Er Ke Za Zhi, 58(8), pp.635–639. doi: 10.3760/cma.j.cn112140-20200426-00437. PMID: 32842383.

